# Prevalence and determinants of opportunistic infections among HIV infected adults after initiation of antiretroviral therapy in Ethiopia: A systematic review and Meta-analysis

**DOI:** 10.1101/2022.10.27.22281601

**Authors:** Beshada Zerfu Woldegeorgis, Zewdineh Zekarias, Bulcha Guye Adem, Mohammed Suleiman Obsa

**Author notes:** **Email address:** Beshada Zerfu Woldegeorgis.

## Abstract

**Background:** Reliable data on the burden of opportunistic infections (OIs) after antiretroviral therapy (ART) initiation is critical for planning health services and reducing OI-related morbidity and mortality. Nevertheless, there has been no nationally representative information on the prevalence of OIs in our country. Therefore, we have undertaken this comprehensive systematic review and meta-analysis to estimate the pooled prevalence, and identify potential risk factors associated with the development of OIs in HIV (Human Immunodeficiency Virus)-infected adults receiving ART in Ethiopia.

**Methods:** Articles were searched in international electronic databases. A standardized Microsoft Excel spreadsheet and STATA software version 16 were used for data extraction and analysis, respectively. The Preferred Reporting Items for Systematic Reviews and Meta-Analysis (PRISMA) checklist was used to write this report. The random-effect meta-analysis model was used to estimate pooled effect. Statistical heterogeneity of the meta-analysis was checked. Subgroup and the sensitivity analyses were also performed. Publication bias was examined funnel plots and the nonparametric rank correlation test of Begg and the regression-based test of Egger. Association was expressed through a pooled odds ratio (OR) with a 95% Confidence Interval (CI)

**Results:** A total of 12 studies with 6163 study participants were included. The overall estimated pooled prevalence of OIs was 43.97% (95 % CI (38.59, 49.34). Poor level of adherence to ART (OR, 5.90 (95% CI (3.05, 11.40), under nutrition (OR, 3.70 (95% CI (2.01, 6.80), CD4 T lymphocytes count <200 cells /µL (OR, 3.23 95% CI (2.06, 5.07), and advanced World Health Organization (WHO) HIV clinical stages (OR, 4.84 (95% CI (1.83, 12.82) were predictors of OIs.

**Conclusion:** The pooled prevalence OIs among adults taking ART is high. Poor adherence to ART, under nutrition, CD4 T lymphocyte counts <200 cells /µL, and presentation with advanced WHO HIV clinical stages were factors associated with the development of OIs.

## Background

Recent global estimates suggest that 79.3 million people have become infected with HIV since the first evidence of the epidemic. Besides, 36.3 million people have died from acquired immune deficiency syndrome (AIDS) related illnesses. In 2020, an estimated 37.7 million people were living with HIV(PLHIV), 1.5 million were newly infected with HIV, 680 000 died from AIDS-related illnesses worldwide, and Sub-Saharan Africa is home to two thirds (67%) of PLHIV [1].

People living with HIV are frequently exposed to co-infections during the course of the disease [2] more frequently and severely, causing significant morbidity and mortality, and necessitating lifelong ART [3, 4].Thus, OIs remain the major driver of HIV-associated morbidity and mortality, accounting for the substantially higher mortality observed in Low and middle-income countries (LMICs)[5]. High levels of early morbidity and mortality following ART initiation continue to be a distinctive feature of ART programs in Sub-Saharan Africa, despite the fact that the introduction and scaling-up of ART have decreased overall mortality [6, 7]

Opportunistic infections are the leading cause of hospitalization and death in patients with HIV and still present formidable challenges for meager healthcare systems endeavoring to provide effective and efficient HIV care [8, 9].In resource-poor settings, between 20% and 52% of hospital beds are occupied by HIV-related OIs [10]. Furthermore, 90% of HIV-related morbidity and mortality are attributed to OIs [11, 12]. Between 2000 and 2019, 170.79 billion United States dollar in development assistance for health was spent on HIV globally and most of this aid went towards care and treatment for HIV [13].

Since the advent of ART, a drastic decrease in the magnitude OIs were observed, but still the burden in industrialized countries differ markedly from those countries in Africa [14]. Though there have been no representative studies reporting the magnitude of OIs in adults in Africa, a systematic review and meta-analysis by MR, B.L., et al. revealed that bacterial pneumonia (32.51%), oral and esophageal candidiasis (24.77%), and bacteremia (23.18%) were the most common infections among HIV-infected children receiving ART in LMICs [15].

In Owerri, Imo State, South East Nigeria, among adults who were receiving ART, the overall prevalence of OIs was 22.4%, with candidiasis (8.6%),tuberculosis(7.7%), dermatitis (5.6%), and chronic diarrhea among the most prevalent cases [16]. Similarly research conducted in seven provinces in Indonesia revealed that OIs were prevalent in 33.51% of the PLHIV, with tuberculosis(48.6%), candidiasis(41.2 %), and diarrhea(20%) being the most common OIs [17]. In Ethiopia, health facility reports showed that OIs prevalence was 88.4% in Tercha hospital, southern Ethiopia [18] and 19.7% in Gonder University hospital, northern Ethiopia [19].

Despite the fact that ART has been proven to be impactful in halting immune system impairment and preventing disease progression, studies showed that OIs have not gone [20, 21] either due to the unmasking of subclinical infection that occurs with immune recovery, drug toxicities and interactions, initial acquisition of a drug-resistant strain or high exposure to infectious agents[22, 23].

A study in the United Kingdom reported that late presentation to health facilities increases the odds of developing OIs and remains a significant problem in developed countries, with over 20% of patients in the United kingdom suffering from OIs [24]. Besides, baseline Neverapine-based regimens, higher viral load, treatment failure, and hemoglobin levels were factors associated with the development of OIs among adult patients taking ART [18, 25, 26]. Furthermore, adherence to ART, nutritional status, isoniazid preventive therapy and cotrimoxazole preventive therapy, place of residence, functional status, gender, age of the PLHIV, CD4 T lymphocytes count and disclosure status have been associated with the development of OIs among adult PLHIV after initiation of ART [26-29].

Reliable data on the burden of OIs after ART initiation is critical for planning health services and reducing OI-related morbidity and mortality. Nevertheless, there has been no nationally representative information on the prevalence of OIs in our country. Therefore, we have undertaken this comprehensive systematic review and meta-analysis to estimate the pooled prevalence, and identify potential risk factors associated with the development of OIs in HIV-infected adults receiving ART in Ethiopia. Results obtained from this review are important for planning delivery of HIV services in Ethiopia which includes the prevention and management of OIs, and provision of comprehensive and high-quality care to HIV patients.

## Methods

### Study Protocol registration and reporting

This systematic review and meta-analysis was undertaken to estimate the prevalence of OIs in HIV-infected adults receiving ART in Ethiopia and identify the associated factors. The study protocol for this review has been registered in an international database, the Prospective Register of Systematic Reviews, by the University of York Centre for Reviews and Dissemination with identification number CRD42022296126 (URLhttps://www.crd.york.ac.uk/prospero/#myprospero).

Furthermore, modifications to the title and expected date of review finalization were made on October 17, 2022. A 17-item Preferred Reporting Items for Systematic Reviews and Meta-Analyses Protocols 2015 checklist was used to guide protocol development [30]. Besides, the Preferred Reporting Items for Systematic Reviews and Meta-Analyses 2020 Checklist was used to report the review’s findings [31](Additional file 1)

### Research questions

We have undertaken this comprehensive systematic review and meta-analysis to seek an answer to the following questions: 1) What is the national burden of OIs in adult PLHIV following the initiation of ART? 2) What are the potential risk factors that affect the occurrence of OIs in adult PLHIV following the initiation of ART?

### Inclusion Criteria

Inclusion criteria for this review were based on the study characteristics and report characteristics determined by using the CoCoPop (condition, context, and population) mnemonic[32]. Thus, we included all observational studies (cross-sectional studies, case-control studies, and cohort studies) in this systematic review and meta-analysis.

#### Participants/Population

The individuals aged 15 and above years who participated in the studies that assessed the prevalence of OIs and/or associated factors were considered. According to Ethiopian national guide lines for HIV/AIDS treatment programs, individuals aged greater or equal to 15 years are considered adults [22].

#### Context

Limited to primary studies conducted in Ethiopia.

#### Language of publication

Articles reported in English were included.

#### Years of publication

Articles published between 2010 and 2022 were included.

### Exclusion Criteria

Studies reporting the outcome of interest in pre-ART PLHIV, without full-text access; articles that contained insufficient information; findings from personal opinions; articles reported outside the scope of the outcome of interest; qualitative study design; case reports; case series; letters; unpublished data; and previous systematic reviews were filtered out.

### Information Sources and Search Strategy

Literature search strategies were developed using medical subject headings (MeSH) and text words related to the outcomes of the study. The search typically included the following electronic bibliographic databases: Excerpta Medica database (EMBASE), PubMed, Web of Science, African Journal of Online (AJOL), Google Schoolar,and Cochrane Library to ensure complete coverage of the topic by accounting for variability between the indexing in each database. The literature search was limited to studies published in English language between 1^st^ January, 2010 and February 30^th,^ 2022 which explored prevalence, and/or factors associated with OIs among adults after initiation of ART. The reference lists of included studies identified through the search were scanned to ensure literature saturation. Where necessary, we also searched the authors’ files to ensure that all relevant materials had been captured.

We reviewed grey literature via Google. In addition, unpublished studies were sought from the official repositories of Ethiopian universities in particular Addis Ababa University and Jimma University. For the advanced search in PubMed, the following steps comprised the search process: Initially, the search statement was divided into main concepts: prevalence, Incidence, Opportunistic infections, associated factors, adults, antiretroviral therapy, and Ethiopia. Subsequently, we gathered keywords from Google scholar, Wikipedia, and Google for each concept, which were then searched independently in PubMed to find MeSH terms in the MeSH hierarchy tree and then combined in an advanced search.

Boolean operators (AND and OR) were used to combine these concepts as follows: (((((((((((Prevalence [Text Word]) OR (“Prevalence”[Mesh] AND (opportunistic infections[Text Word])) OR “AIDS-Related Opportunistic Infections”[Mesh])) AND (HIV[Text Word])) OR (“hiv”[MeSH Terms])) AND Predictors[Text Word] OR (Risk Factors[Text Word]))OR (“Associated factors”[Text Word] OR (“Risk Factors”[Mesh])) AND (adults[Text Word])) OR (“adult”[MeSH Terms])) “anti-retroviral agents”[MeSH Terms] OR antiretroviral [Text Word] AND (Ethiopia (text word))) OR (“Ethiopia*”[Mesh]). Finally, we filtered the results to include just the most relevant ones. The search was double-blinded and conducted from February 30^th^ to April 10, 2022, by two authors (**BZ.W and MS.O**). A separate file with the search details was supplied (Additional file 2)

### Study Selection Procedures

The articles that were found through the electronic database searches were exported to the reference management software, EndNote X7, where duplicate studies were then eliminated. Two authors (**BZ.W and MS.O**) independently screened the titles and abstracts that were obtained by the search against the inclusion criteria. To describe the extent to which assessments by multiple authors are similar, inter-rater agreement was calculated after referring to the Cochrane handbook for systematic reviews.

In this case, kappa greater or equals 75% was considered, indicating excellent agreement. The screened articles were then subjected to a full article review by two independent authors (**ZZ and BG.A**). A pre-defined eligibility criterion was used to determine which records were relevant and should be included in the review. Where more information was required to answer queries regarding eligibility, the remaining authors were involved. Disagreements were resolved through discussion. Moreover, the reasons for excluding the articles were recorded at each step.

### Data Extraction

Two authors (**BZ.W and MS.O**) abstracted the relevant data independently by using a standardized Microsoft Excel spread sheet. For data extraction, Joana Briggs Institute (JBI) - adopted formats were employed [33].The first author’s name, sample characteristics, regions of study, year of publication, study design, study area, outcome measures, timing and procedures of data collection, response rates, knowledge of and attitudes toward epilepsy were collected.

The reliability agreement among the data extractors was evaluated and verified using Cohan’s kappa coefficient after data was recovered from 30 percent of the primary studies. As a consequence, the kappa coefficient’s strength of agreement was divided into four categories: low (0.20), fair (0.21-0.40), moderate (0.41-0.60), good (0.61-0.80), and virtually perfect agreement (0.81-1) [34].A kappa statistic value of more than or equal to 0.5 was regarded as congruent and acceptable. In the case of disagreements between the two data extractors, a third author (**BG.A)** was involved in adjudicating unresolved disagreements through discussion and re-checking of the original articles.

### Outcome measurement

The number of adult patients who had OIs was divided by the total number of adults taking ART and multiplied by one hundred to calculate the pooled prevalence of OIs among adults after the initiation of ART. The pooled effect for associated factors was investigated using the OR. Furthermore, variables identified as a risk factor for OIs in at least three studies were taken into account.

### Definition of terms

Authors of primary studies classified adherence to ART as good, fair, or poor using the definition from the national HIV care follow-up form. Good adherence was defined as taking more than 95% of prescribed doses, fair adherence as taking 85–94% of prescribed doses, and poor adherence as taking less than 85% of prescribed doses as documented in patient records. Nutritional status was also based on national HIV care and treatment guidelines, and a body mass index of less than 18.5kg/m2 was a cut-off point for under nutrition irrespective of its severity. Furthermore, for adolescents and adults, advanced HIV diseases were defined as CD4 cell counts of 200 cells/mm3 or a WHO stage III or IV event [22]

### Methodological Quality (Risk of Bias) Assessment

To assess the quality of the studies, the Joana Briggs Institute (JBI) critical appraisal checklists for descriptive cross-sectional studies comprised of 8 components (Additional file_ 3A), analytic cross-sectional comprised of 7 components (Additional file_3B), cohort studies comprised of 11 components (Additional file_3C), and case control studies comprised of 10 components (Additional file_3D) were employed. Three authors (**ZZ, BG.A and MS.O**) independently assessed the methodological quality of each study. Disagreements were resolved through consultation with a third independent reviewer (**BZ.W**). Studies with a score of 7 or higher after being evaluated against these criteria were considered low risk and included in this systematic review and meta-analysis.

### Data synthesis and Meta-Analysis

The extracted data were imported from a Microsoft Excel spreadsheet into STATA MP 16 statistical software (StataCorp LP, 4905 Lakeway Drive, College Station, TX 7845, USA) for analysis. The heterogeneity of the results was visually examined via the forest plots with pooled estimates. Thus, its presence was confirmed subjectively with a lack of overlap between the confidence interval (CI).

In additions, the statistical heterogeneity was explored more formally by using Cochran’s Q test (x^2^) at P-value < 0.10 indicating significant heterogeneity. Another heterogeneity measure, Higgins and Thompson’s I^2^ statistics, was employed to estimate the percentages of the between-study variability where, 0%, 25 to50%,50 to 75%,and greater than or equals to75% indicated no heterogeneity, low heterogeneity, moderate heterogeneity, and high heterogeneity respectively[30].

The random-effect (RE) meta-analysis model was used to estimate Der Simonian and Laird’s pooled effect due to the presence of a considerable statistical heterogeneity. Subgroup meta-analysis based on the study regions as covariates, meta regression, and sensitivity analyses were also performed to investigate the source of statistical heterogeneity. Publication or dissemination bias was examined subjectively using funnel plots and objectively using the nonparametric rank correlation test of Begg[35] and the regression-based test of Egger for small study effects [36], with P <0.05 being taken into consideration to declare potential publication bias. Results were presented in the form of tables, texts, and figures

## Result

### Search and Study Selection

Our search was restricted to articles published in English language between 1^st^ January, 2010 and February 30^th,^ 2022 in the electronic databases of PubMed, Web of Science, and EMBASE. In addition, Google, Google scholar, and AJOL were searched. Through systematic and manual searching, 640 primary articles were found. Due to duplication, 572 articles were removed. The remaining 68 were screened based on their title and abstract, with 38 being eliminated as unrelated to our study. Finally, 30 full-text primary articles were evaluated against eligibility criteria, and 16 were selected for quantitative analysis (**Fig 1**).

**Fig 1.**
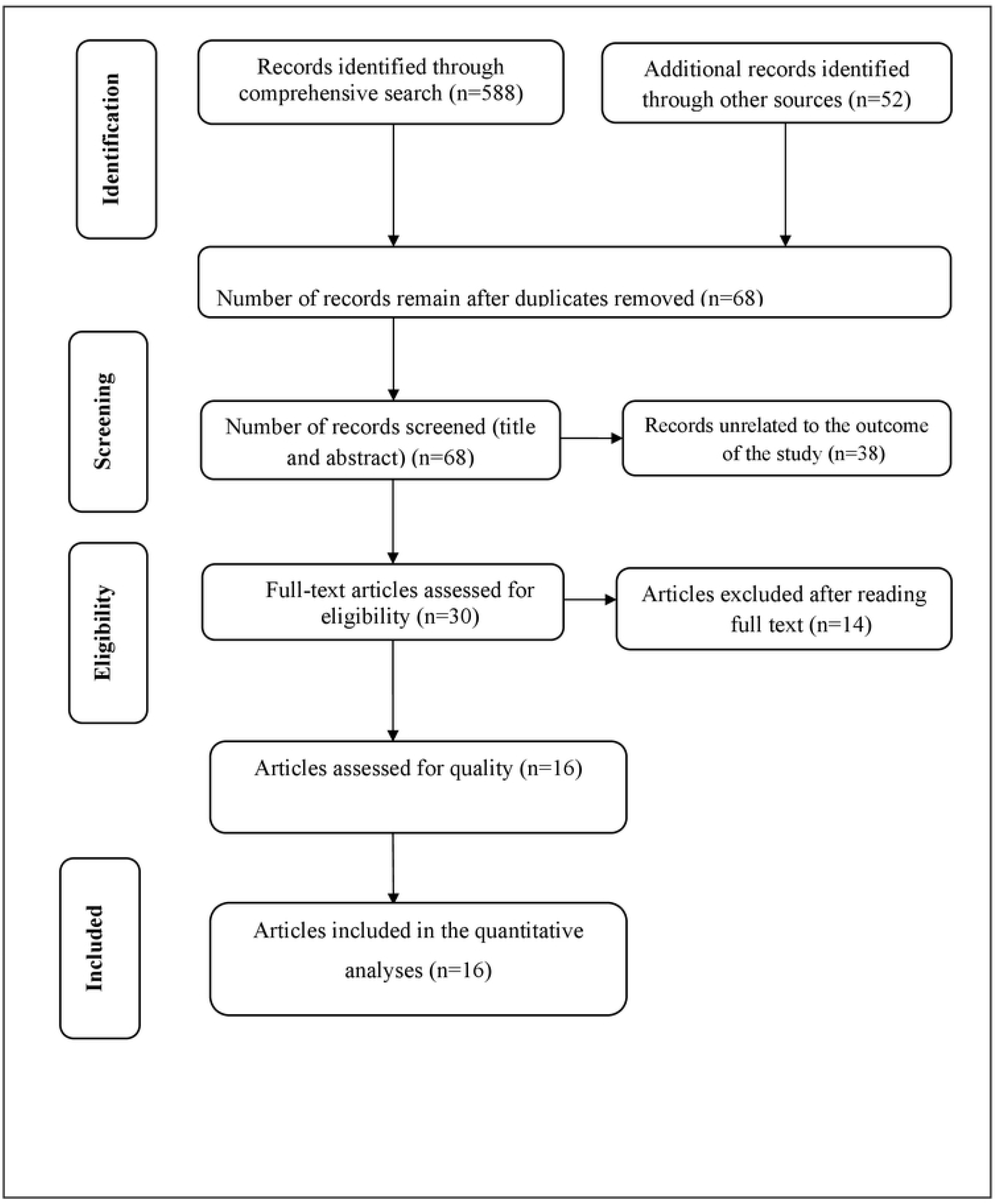
PRISMA flow diagram of included studies in the systematic review and meta-analysis.

### Study characteristics

This systematic review and meta-analysis included 16 studies involving 6163 participants, with a response rate ranging from 96% to 100%. More than 50% (n= 3558) of the participants were female. Thirteen studies employed a cross-sectional study design [18, 19, 37-47]. The remaining two were cohort studies[26, 28], and one was a case control study[48]. All studies were conducted from 2013 to 2021 in different geographic locations of Ethiopia. Four of the primary studies included were based in Ethiopia’s Southern Nations Nationalities Peoples’ Regional States (SNNPRs) [18, 28, 42, 44], three in Amhara[19, 45, 48], three in Addis Ababa [37, 38, 43], three in Tigrai [26, 41, 46], one in Oromia [47], one in Sidama [39], and one in Harari [40].The highest (88.44%) [18] and the lowest (19.72%)[19] Prevalence of OIs was reported in studies conducted in Tercha Hospital, southern Ethiopia, and Gonder University Hospital, northern Ethiopia, respectively (**Table 1**).

**Table 1:**
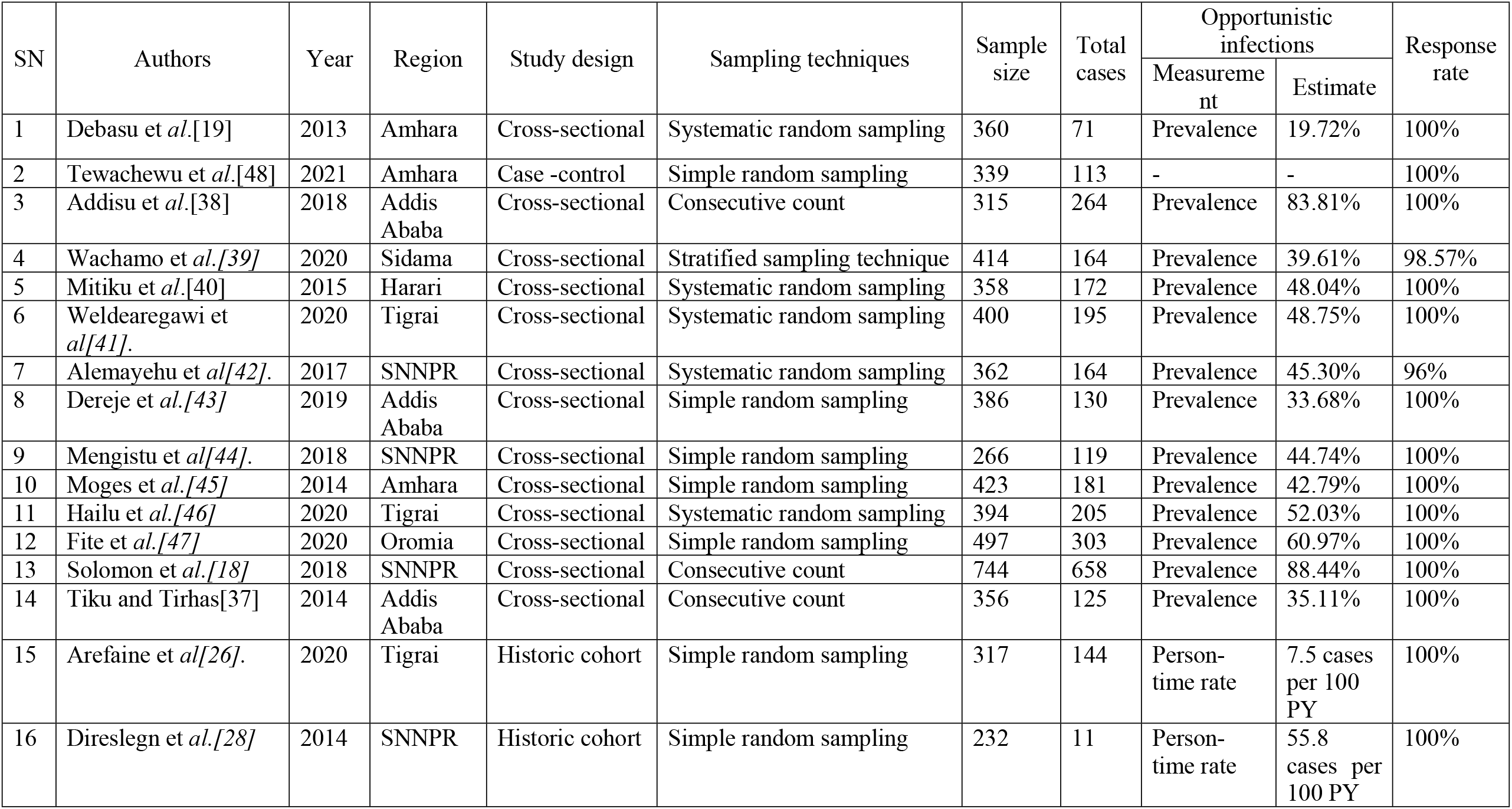
The characteristics of the studies included in the systematic review and meta-analysis.

Furthermore, authors of 13 primary studies assessed the spectrum of OIs [18, 19, 26, 37-45, 47]. In terms of sampling techniques, systematic random sampling techniques [19, 40-42], simple random sampling techniques [26, 28, 43-45, 47, 48], combined stratified and simple sampling techniques[39] and consecutive count [18, 37, 38] were used to select the required study subjects. Furthermore, the authors of these studies employed a structured questionnaire, and or a pre tested and refined checklists to obtain necessary data. A complete summary of descriptive characteristics of all included primary studies are provided (see **Additional file 4**).

### Prevalence of opportunistic infections

Given the substantial statistical heterogeneity in the fixed-effects model, the pooled estimate was determined using a random effects model. Thus, an overall pooled prevalence of OIs among HIV infected adult patients on ART in Ethiopia was found to be 43.97% [(95 % CI: 38.59, 49.34), I^2^=92.4%, P <0.001, n=4195] (**Fig 2**).

**Fig 2:**
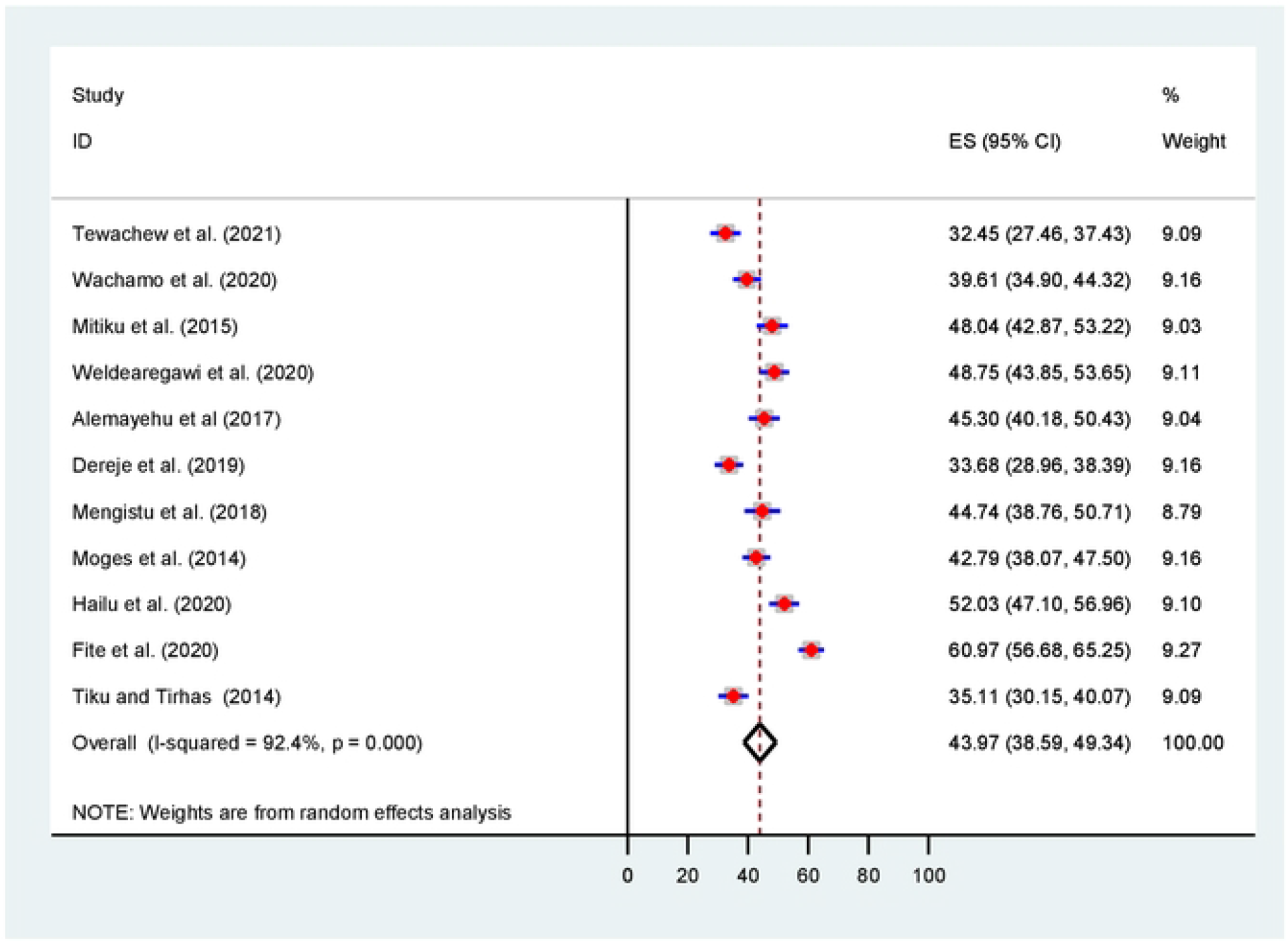
Forest plot for the pooled prevalence of opportunistic infections.

### Subgroup meta-analysis

Subgroup analysis was conducted based on Ethiopia’s administrative regions where primary studies were based, and year of publication. Thus, we observed regional variations in the prevalence of OIs in this review. The prevalence of OIs was found to range between 31.36% [(95% CI: 30.94, 37.78), I^2^=0.0%, P<0.0681, n=762] in random effects pooled meta-analysis in Amhara region and 60.97 %(n=497) in Ethiopia’s Oromia region with single study (**Fig 3**).

**Fig 3:**
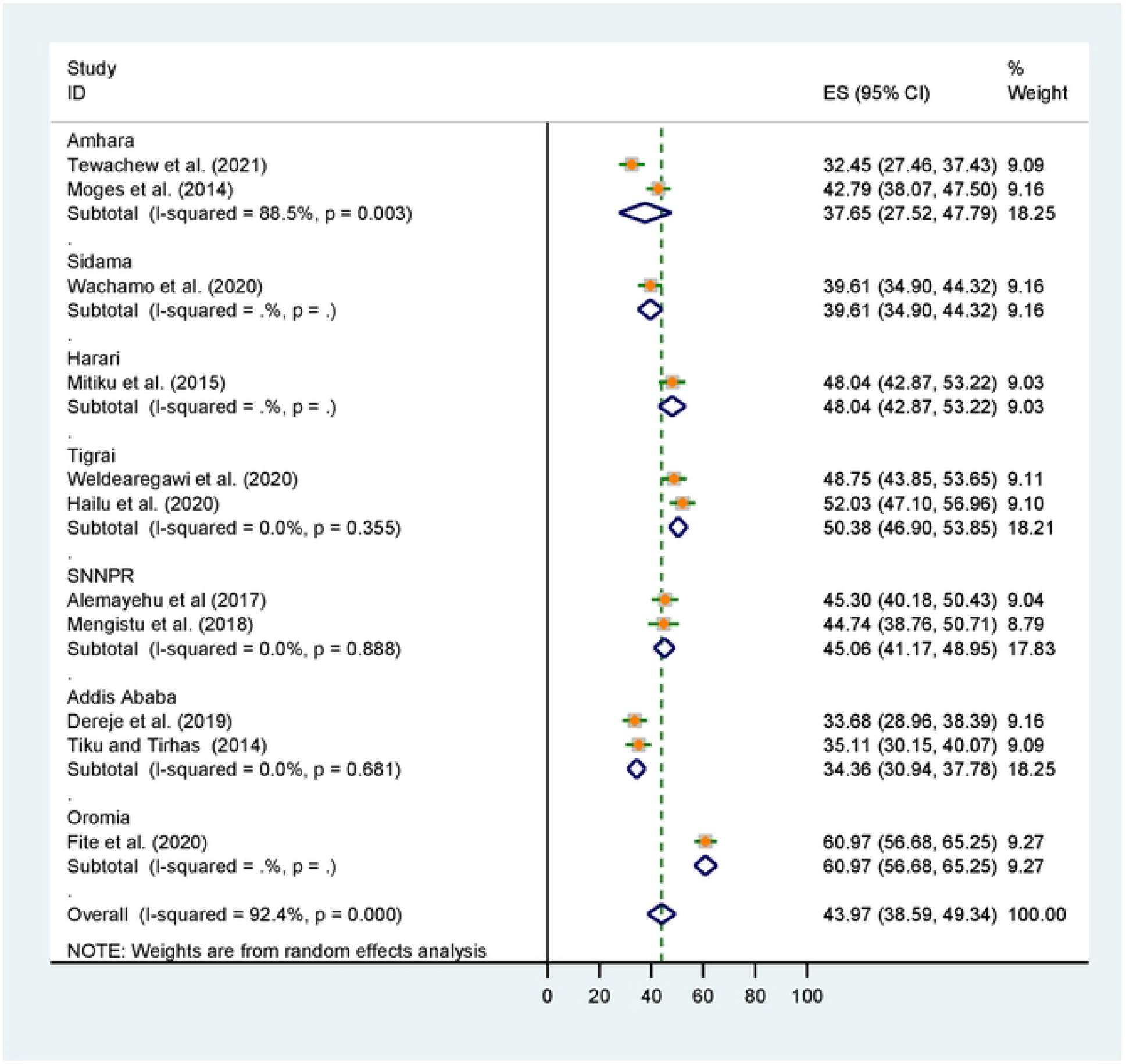
Subgroup analysis for pooled prevalence of OIs by Ethiopia’s region.

Considering the introduction of WHO test and treat recommendation, we also conducted subgroup analysis by categorizing in to before and after 2016. Thus, the pooled prevalence of OIs after the test and treat recommendation was found to be 44.71% [(95% CI: 37.68, 51.75), I^2^=93.9 %, P<0.001, n=3058] (**Fig 4**).

**Fig 4:**
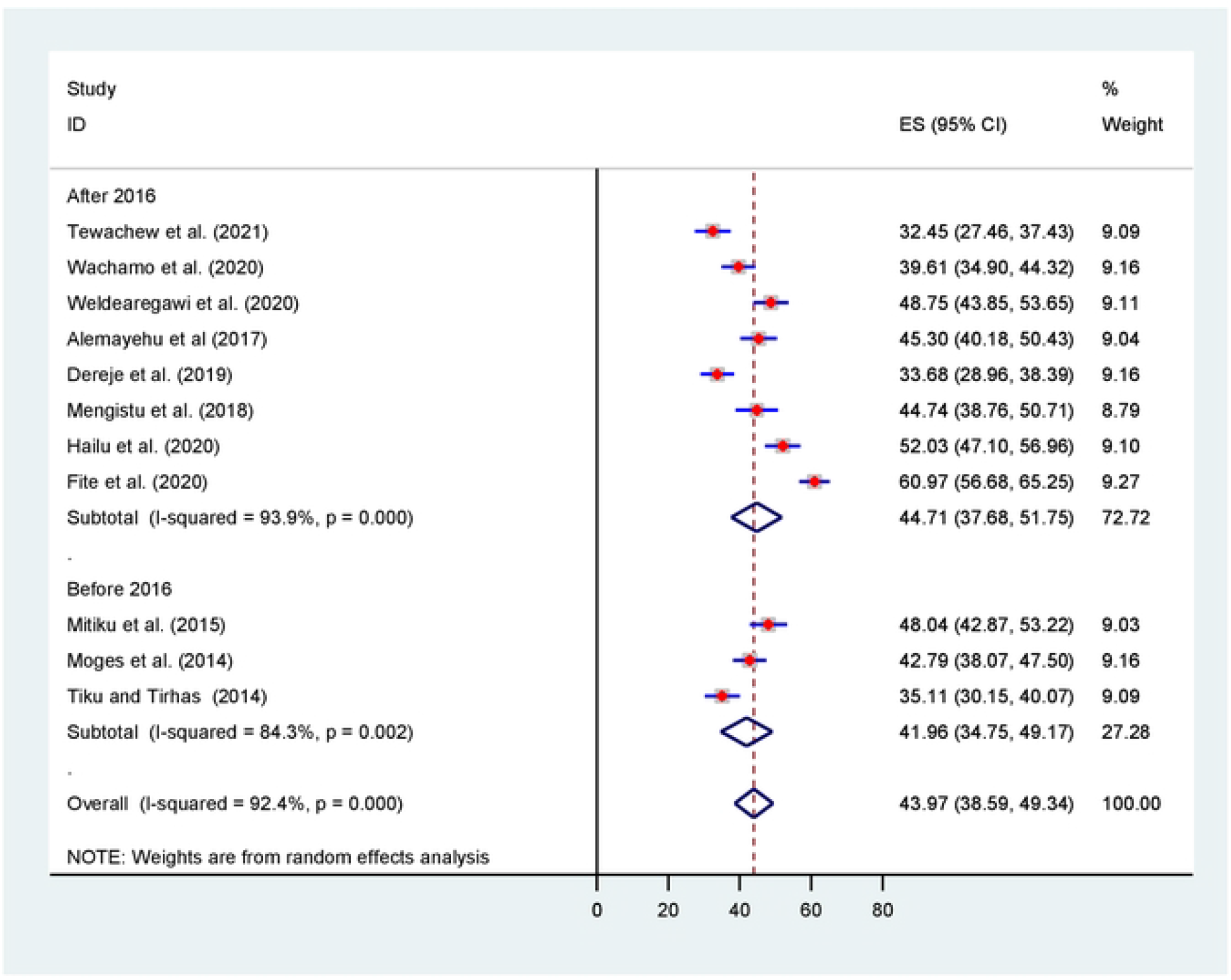
Subgroups analysis for prevalence of OIs by year of publication.

### Meta-regression

Random-effects Meta-regression using sample size and year of publication as covariates was performed to explore the source of heterogeneity at a 5% significance level. As shown in **Table 2**, these covariates were not found to be the source of heterogeneity.

**Table 2:** Meta regression analysis of factors affecting between study heterogeneity.

| Outcome | Source of heterogeneity | Coefficient | Standard error | t | P>\|t\| | [95% confidence interval] |  |
| --- | --- | --- | --- | --- | --- | --- | --- |
| Prevalence of opportunistic infections | Year of publication | .0021707 | .3074093 | 0.01 | 0.995 | -.7067165 | .7110578 |
|  | Sample size | .0019004 | .0141767 | 0.13 | 0.897 | -.0307911 | .0345919 |
|  | -Cons | -1.348413 | 619.2611 | -0.00 | 0.998 | -1429.367 | 1426.67 |

### Sensitivity meta-analyses

We conducted a leave-out-one meta-analysis to explore the effect of each study on the overall prevalence of OIs while gradually excluding each study. Results showed that the combined prevalence of OIs did not significantly change as a result of the excluded study (**Table 3**).

**Table 3:**
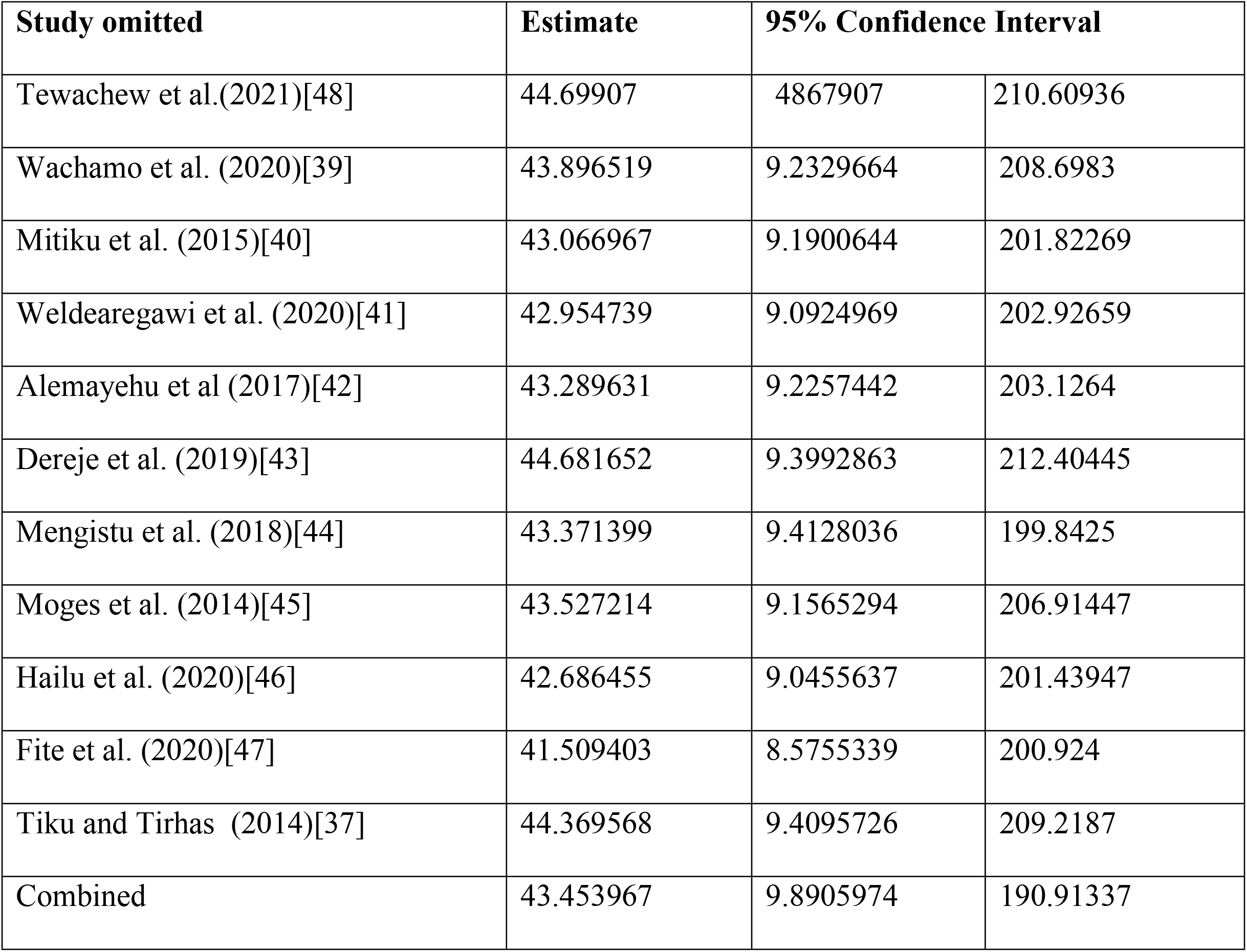
Sensitivity analysis of pooled prevalence with each study removed one by one.

**Table 4:** Factors associated with development of OIs among adults after initiation of ART in Ethiopia.

| **Variables** | **Comparison** | **Number of studies** | **Sample size** | **Pooled OR(95% CI)** | **P.value** | **I<sup>2</sup>(%)** | **Heterogeneity test** | **Egger test** |
| --- | --- | --- | --- | --- | --- | --- | --- | --- |
| Adherence to ART [18, 26, 45, 46, 48] | Poor vs. fair and good | 5 | 2205 | 5.90(3.05,11.40) | <0.001* | 76.4 | 0.002 | 0.881 |
| Under nutrition [44, 45, 48] | Yes vs.No | 3 | 1029 | 3.70(2.01,6.80) | <0.001* | 71.4 | 0.030 | 0.753 |
| CD4+ T lymphocyte count(cells/μL) [18, 19, 38-40, 44, 46] | <200 vs.≥200 | 7 | 2848 | 3.23(2.06,5.07) | <0.001* | 79.8 | <0.001 | 0.169 |
| Functional status [26, 40, 44, 46] | Bedridden vs. ambulatory and working | 4 | 1332 | 2.41(0.56,10.46) | 0.239 | 93.4 | <0.001 | 0.165 |
| Hemoglobin(gm/dl) [18, 26, 45] | <10 vs.≥10 | 3 | 1626 | 1.49(0.46,4.82) | 0.505 | 92.3 | <0.001 | 0.206 |
| Advanced WHO HIV clinical stage [19, 40, 43-45] | III and IV vs. I and II | 5 | 1599 | 6.75(3.87,11.76) | <0.001* | 77.6 | 0.001 | 0.566 |

*\*-Statistically significant at $P<0.05$*

### Publication bias (Reporting bias)

Publication bias was assessed subjectively using a funnel plot and objectively by the regression-based test of Egger and the nonparametric rank correlation test of Begg at P<0.05. A funnel plot showed symmetrical distribution (**Fig 5)**.

**Fig 5:**
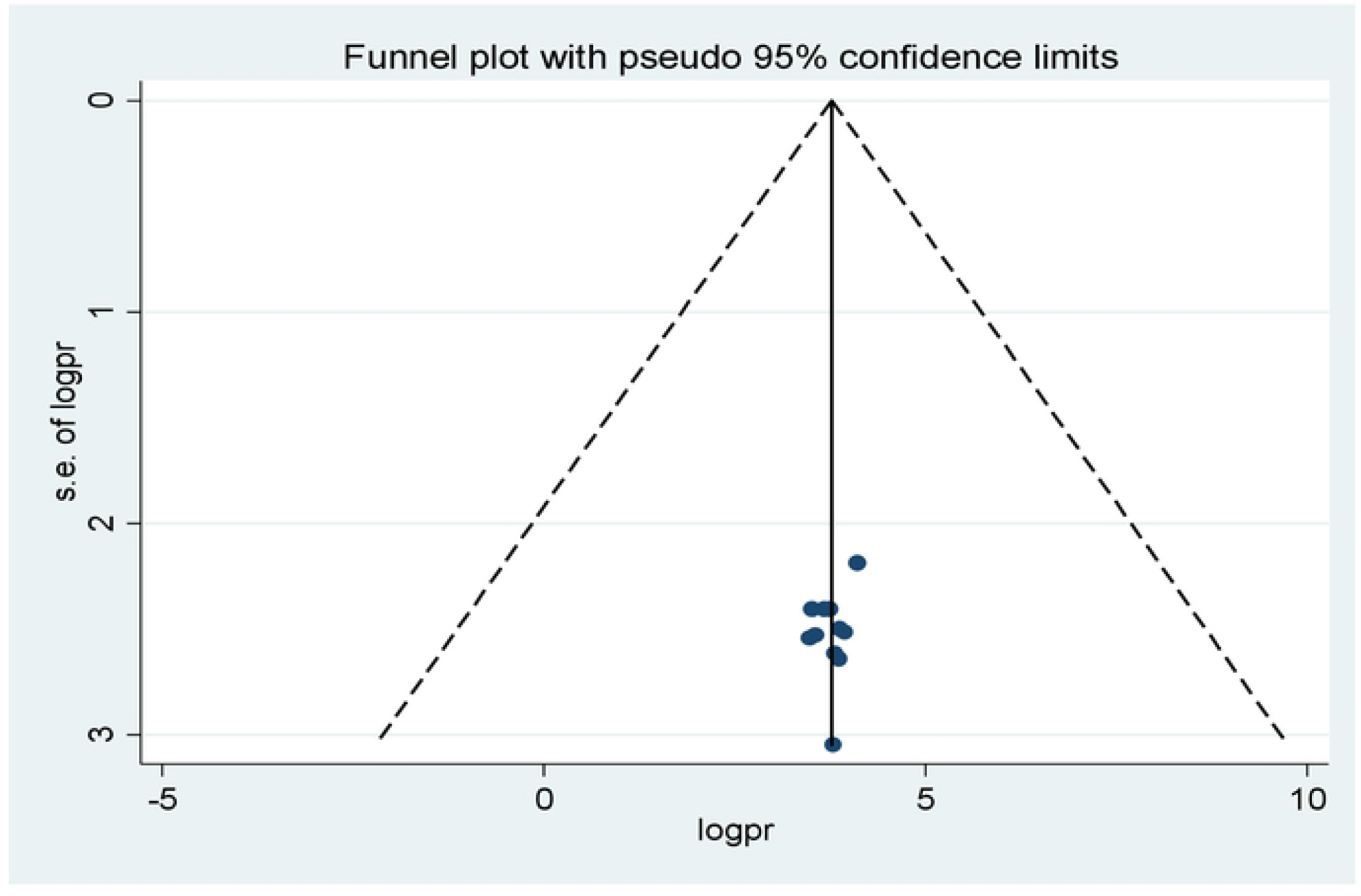
Funnel plots for exploring publication bias.

Moreover, neither Egger’s linear regression test (t = −0.67, P = 0.517) nor Begg’s rank correlation test (z = 0.16, P = 0.876) were statistically significant for a pooled prevalence of opportunistic infections, corroborating that there is no evidence of small study effects.

### Spectrum of opportunistic infections

The authors of these primary studies included in this review assessed the patterns of common OIs [18, 19, 26, 37-45, 47]. Accordingly, oral candidiasis (25.31%) was the most frequently occurring OI among HIV-infected adults after the initiation of ART. Pulmonary tuberculosis (22.04%), herpes zoster (14.95%) ranked second and third respectively (**Fig 6**).

**Fig 6:**
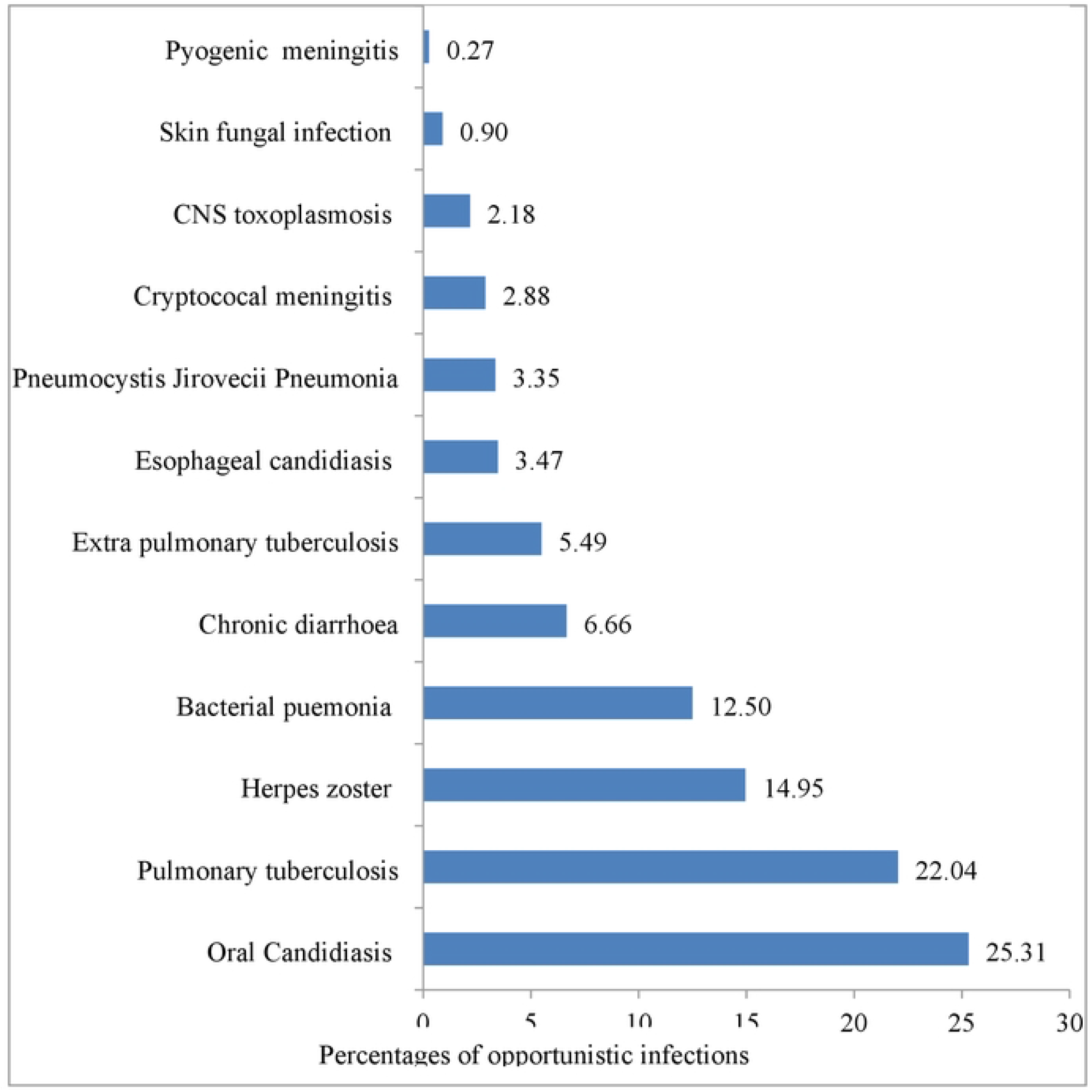
The patterns of common OIs among adults on ART in Ethiopia, 2010-2021.

### Factors associated with the development of opportunistic infections

About fifteen primary studies included in this meta-analysis have reported various factors as determinants of OIs among HIV-infected adult patients who were taking antiretroviral therapy in Ethiopia [18, 19, 26, 28, 37-41, 43-48]. We identified five predictor variables out of seven extracted variables, each reported by at least three primary studies, that were found to be associated with the development of OIs. These include level of adherence to ART, body mass index, baseline CD4 T lymphocytes count, and WHO HIV clinical stage. The pooled OR was used to estimate the association between these variables and occurrence of OIs, and a statistically significant association between the outcome and predictor variables were declared at two –sided P<0.05 with 95 % CI.

To begin with, the odds of developing OIs were approximately 6 times [OR, 5.90 (95% CI (3.05, 11.40), P<0.001, I^2^=76.4%)] higher in adult PLHIV who had good level of adherence to ART compared to fair and good adherents. Additionally, the odds of developing OIs was 3.7 times higher in adult PLHIV with under nutrition than in adult PLHIV with normal body mass index [OR, 3.70 (95% CI (2.01, 6.80), P<0.001, I^2^=71.4%)]. In this meta-analysis it was also found that the odds of OIs occurring was 3.23 times [OR, 3.23 95% CI (2.06, 5.07),P<0.001,I2=79.8%)] more likely in PLHIV who had baseline CD4 T lymphocytes less than 200 cells per microliter compared to their counter parts. Moreover, the odds of exhibiting OIs were approximately 5 times [OR, 4.84 (95% CI (1.83, 12.82), P=0.002, I2=89.5%)] more likely in adult PLHIV who had presented with advanced WHO HIV clinical stage at enrollment to the program.

## Discussion

In this systematic review and meta-analysis, we sought to estimate the pooled prevalence of OIs and identify potential predictors among adult PLHIV after initiation of ART in Ethiopia. The overall random effects meta-analysis revealed that the pooled prevalence of OIs was 43.97% [(95 % CI: 38.59, 49.34). This finding was by far higher than OIs prevalence in Owerri, Imo State, South East Nigeria(22.4%)[16],Indonesia(33.51%)[17], and Uganda (6.7%)[49].This can be explained by study design, and sample size. Another possible explanation could be due to a disparity in the level of engagement in HIV care in these countries.

In terms of spectrum and magnitude, the six most common OIs identified in this review were: oral candidiasis (25.31%), pulmonary tuberculosis (22.04%), herpes zoster (14.95%), bacterial pneumonia (12.50%), chronic diarrhea (6.66%), and extra pulmonary tuberculosis (5.49%). The pattern differs from a previous national cross-sectional study that examined HIV patients’ medical records to determine the prevalence of OIs in selected Ethiopian health facilities over a one-year period (2003-2004)[50]. This could be due to the latter involving both pre-ART and PLHIV receiving ART.

Furthermore, the study period may have an impact on the apparent findings. However, it is congruent with the study in Indonesia [51], where tuberculosis, candidiasis, and chronic diarrhea were the major causes of morbidity. In addition, the pattern is not different from a review finding in LMIC, where Oral candidiasis, unspecified tuberculosis, herpes zoster, pulmonary tuberculosis, and bacterial pneumonia were the most common OIs in ART-naive participants [52].

In this analysis, we also found that poor adherence to ART, malnutrition, CD4 T lymphocyte counts of less than 200 cells per microliter, and presentation with advanced WHO HIV clinical stages were potential determinants of OIs among adult PLHIV after commencement of ART. To begin with, adult PLHIV who were poorly adherent to ART had a strong risk of developing OIs. This finding was congruent with other study’s findings from a west African country, Nigeria[53]. This is because poor adherence to ART reduces the effectiveness of ARV drugs while also accelerating viral replication and immune suppression, creating a favorable environment for the development of OIs.

Being under nourished was another determinant factor that affects the accusation of OIs among adult PLHIV. This finding was in agreement with previous studies in KwaZulu-Natal, South Africa, where low body mass index was associated with morbidity and mortality among HIV patients [54]. This could be due to a complex and mutually reinforcing relationship between malnutrition and HIV infection. Furthermore, malnutrition can result from HIV-induced immune impairment and the resulting OIs can cause poor appetite and nutrition absorption from the gastrointestinal.

Moreover, adult PLHIV whose baseline CD4 T lymphocytes measured less than 200 cells per microliter had three times higher odds of developing OIs compared to their counterparts. This finding is concordant with a study in India, which was conducted among AIDS patients in Mangalore, Karnataka, and found that patients with CD4 counts of 200 cells/mm3 were at a high risk of developing advanced form of OIs such as tuberculosis, Pneumocystis jiroveci pneumonia, and cryptococcal meningitis [55]. Another study by Ghate et al., carried out to identify OIs among HIV-infected individuals by stages of immunodeficiency, revealed that PLHIV with baseline CD4 counts of less than 200 cells per microliter were six times more likely to develop OIs [56]. This finding appears to be correct because CD4 T lymphocytes play detrimental role in the induction of both humoral and cellular immunity to combat OIs.

In this review, baseline advanced (III and IV) WHO HIV clinical stage was another potential determinant of OIs after the initiation of ART than stage I and II. Our finding was consistent with a report from Mulago-Mbarara teaching hospitals’ joint AIDS program, Kampala, Uganda[49],Nigeria[57],and South African cohort[58].This can be explained by the fact that the progression of HIV from infection through subtle symptoms to advanced HIV disease is correlated with prolonged immune suppression, which is partly a reflection of late presentation to health facilities. Besides, individuals with advanced HIV diseases are prone to acquiring multiple other OIs.

### Limitations and strengths of this study

The limitations of this systematic review and meta-analysis need to be acknowledged. To begin with, primary studies from Ethiopia’s Oromia region, which has the most health care facilities providing HIV care and treatment, are underrepresented in the pooled random effect analysis. Nonetheless, HIV care and treatment in Ethiopia is a vertical program that is consistent with other regions of the country. Furthermore, because there has been no previous nationally representative study, our findings are difficult to compare. Therefore, the results need to be interpreted with caution.

On the other hand, the current systematic review and meta-analysis has also strengths. The outcome measurement across all studies was consistent, which could be due to the HIV care program being based on national guidelines, which are available in all health facilities. Besides, observational analytic studies were involved, so it was possible to infer a causal effect relationship. The protocol for this study has been registered. More than seven online databases were searched to avoid missing published studies, including articles published in African journals. In addition, a manual search was performed to retrieve the article using Google Scholar. During the selection of articles, the Preferred Reporting Items for Systematic Reviews and Meta-Analyses 2020 Checklist were strictly followed, and the articles were closely assessed for their quality using the newly amended JBI critical appraisal tool. Furthermore, we used broader inclusion criteria to ensure we did not miss articles.

### Conclusion and recommendations

In summary, the pooled random effect meta-analysis revealed that the percentage of OIs among adult PLHIV after the commencement of ART is high. In the subgroup meta-analysis, regional variations in the prevalence were observed, with the highest in the Oromia region and the lowest in the Amhara region. Oral candidiasis, pulmonary tuberculosis, herpes zoster, bacterial pneumonia, chronic diarrhea, and extra pulmonary tuberculosis are the six major OIs. Moreover, poor adherence to ART, under nutrition, CD4 T lymphocyte counts of less than 200 cells per microliter, and presentation with advanced WHO HIV clinical stages were associated with the development of OIs.

Therefore, it is critical to provide intensified care for patients presenting with advanced HIV disease, lower immunologic status and under nutrition which include screening prompt provision of chemoprophylaxis in order to reduce or avoid the risk of developing OIs after ART initiation. Furthermore, enhanced adherence counseling should be provided based on the patient’s adherence track records.

## Data Availability

All relevant data are within the manuscript and its Supporting Information files

## Availability of data and materials

All data about this study are contained and presented in this document

## Abbreviations/Acronyms

AIDS: Acquired Immunodeficiency Syndrome
ART: Antiretroviral Therapy
CI: Confidence Interval
HIV: Human Immunodeficiency Virus
JBI: Johanna Briggs Institute
LMICs: Low-and Middle Income countries
OIs: Opportunistic Infections
OR: Odds ratio
PLHIV: People Living with HIV
SNNPRs: Southern Nations Nationalities Peoples’ Regional State
WHO: World Health Organization

## Acknowledgements

We would like to thank the authors of the primary studies included in this systematic review and meta-analysis

## Author contributions Statement

BZW conceptualized the study, prepared and registered study protocol. MSO, BZ.W, BGA and ZZ searched, screened articles and involved in the risk of bias assessment. MSO and BZW were involved in data abstraction, statistical analysis results interpretation, and writing the initial and final drafts of the manuscript. All authors proofread and approved the final manuscript.

## Funding

The authors received no specific funding for this work

## Ethics approval and consent to participate

Not applicable

## Consent for publication

Not applicable

## Competing interests

We declare no computing interest

## Notes

### Competing Interest Statement

The authors have declared no competing interest.

### Funding Statement

The author(s) received no specific funding for this work

